# Digital Spatial Profiling identifies phospho-JNK as a biomarker for early risk stratification of aggressive prostate cancer

**DOI:** 10.1101/2024.12.20.24319439

**Authors:** Samaneh Eickelschulte, Adam Kaczorowski, Florian Janke, Anja Lisa Riediger, Olga Lazareva, Sarah Böning, Glen Kristiansen, Constantin Schwab, Albrecht Stenzinger, Holger Sültmann, Stefan Duensing, Anette Duensing, Magdalena Görtz

## Abstract

**Background:** Prostate cancer (PCa) is a highly heterogeneous disease, with cases ranging from indolent to highly aggressive forms. Ongoing research focuses on identifying new biomarkers to improve early risk stratification in PCa, addressing current limitations to accurately evaluate disease progression. A promising new approach to aid PCa risk stratification is digital spatial profiling (DSP) of PCa tissue.

**Methods:** A total of 94 regions of interest from 38 PCa patients at first diagnosis were analyzed for the expression of 44 proteins, including components of the PI3K-AKT, MAPK, and cell death signaling pathways as well as immune cell markers. An additional validation cohort consisting of 154 PCa patients with long-term follow-up data was analyzed using immunohistochemistry (IHC) to assess the consistency of the identified biomarkers across a larger sample set.

**Results:** DSP identified proliferation marker Ki-67 and phospho-c-Jun N-terminal protein kinase (p-JNK), a member of the MAPK signaling pathway, as significantly upregulated proteins in aggressive PCa (Gleason scores 4 and 5) compared to indolent disease (Gleason score 3). The upregulation of p-JNK was confirmed through IHC. High p-JNK expression was associated with a shorter time to biochemical recurrence (log-rank, p=0.1).

**Conclusion:** Our results indicate that p-JNK may contribute to PCa progression and serve as an early biomarker for aggressive PCa stratification. Identifying this biomarker through DSP could be crucial in advancing disease management and addressing the critical unmet need for more targeted therapies in the treatment of aggressive PCa. Further studies are recommended to evaluate the role of p-JNK in PCa progression.

## 1 Introduction

Prostate cancer (PCa) is the most frequent malignancy in men (1) and is characterized by its high heterogeneity and complexity. In case of elevated prostate-specific antigen (PSA) and/or digital rectal examination (DRE), a prostate biopsy is performed and histopathologically evaluated using the Gleason score grading system. The Gleason score is based on the classification of prostate adenocarcinoma growth patterns, with the separation between Gleason patterns 3 and 4 occurring early in PCa progression (2, 3). The distinction between Gleason 3 and 4 patterns is crucial, as the transition from pattern 3 to pattern 4 indicates a significant shift toward more aggressive disease, which is associated with a higher risk of cancer progression and metastasis (4).

The Gleason score grading system has consistently demonstrated significant prognostic value in stratifying PCa. However, recent research has highlighted the disease as a heterogeneous array of molecular alterations, posing challenges for its diagnosis and treatment (5, 6). Therefore, additional diagnostic methods are often necessary to effectively stratify PCa into high- and low-risk categories, enabling the selection of appropriate therapies for each patient at the time of initial diagnosis. For patients with indolent disease, active surveillance with regular monitoring is the preferred approach, as it avoids unnecessary radical treatment while ensuring timely intervention if the disease progresses (7).

This heterogeneity in PCa is further emphasized by spatial relationships between the cell populations and biomarkers, which can significantly influence disease progression and treatment outcomes (8). Consequently, spatial profiling of protein expression might aid to better understand the biology of PCa and facilitate risk stratification. Biomarkers related to immune cell signaling and cell death pathways hold significant potential for improving PCa prognosis (9-11). Specially, expression of genes related to cell cycle progression, in particular, Ki-67 has been associated with poor prognosis across various types of cancer (12-14), including PCa (15). Additionally, the PI3K/AKT pathway, regulating cell growth, survival, and metabolism, provides valuable insights into the molecular mechanisms that drive aggressive PCa (16). Recent research has also revealed that the complex interactions between the mitogen-activated protein kinase (MAPK) signaling pathway and other cell-signaling cascades can further contribute to PCa progression, highlighting its role in the disease’s development (17). However, the full extent of the MAPK signaling network during prostate tumorigenesis, as well as the involvement of spatial niches in tumor progression and disease recurrence, remain to be fully determined. Studies have shown that c-Jun N-terminal kinase (JNK), a key member of the MAPK signaling pathway, is associated with tumor progression and survival of various cancers (18-20) and contribute to the growth of PCa (21). In this proof-of-concept study, we used digital spatial profiling (DSP) to examine protein expression across four key cellular pathways involved in PCa. We identified Ki-67 and phospho-JNK (p-JNK) as key drivers of disease progression and recurrence distinguishing aggressive Gleason 4 and 5 PCa from indolent Gleason 3 disease.

## 2 Methods

### 2.1 Study population

Tissue microarrays (TMAs) were obtained from localized and locally advanced (N1M0) PCa patients who had received radical prostatectomy at first diagnosis at the Department of Urology, University Hospital of Heidelberg, Germany. The study design comprised specimens from 192 patients, divided into two independent cohorts: one cohort subjected to analysis by DSP (n=38) and a larger validation cohort with long-term follow-up data (n=154) analyzed by immunohistochemistry (IHC) (**Table 1**). All tissue-based experiments in this study were in accordance with the regulations of the tissue bank as well as under the approval of the Ethics Committee of the University of Heidelberg School of Medicine.

**Table 1.** Patient characteristics.

| **Prostate Cancer patients** |  | **DSP cohort (N=38)** | **IHC cohort (N=154)** |
| --- | --- | --- | --- |
| Age, years, median (range) |  | 65 (49–76) | 66 (48–79) |
| PSA level at diagnosis, ng/ml, median (range)<sup>1</sup> |  | 7 (1.52–40) | 0.36 (0.18–1165) |
| ISUP grade group, Gleason score, n (%)<sup>2</sup> |  |  |  |
|  | 2 (7a) | 27 (71) | 38 (25) |
|  | 3 (7b) | 4 (11) | 51 (33) |
|  | 4 (8) | 0 (0) | 60 (39) |
|  | 5 (9–10) | 7 (18) | 4 (3) |
| pT stage, n (%) |  |  |  |
|  | T2 | 22 (58) | 26 (17) |
|  | T3 | 16 (42) | 114 (74) |
|  | T4 | 0 (0) | 14 (9) |
| pN stage, n (%) |  |  |  |
|  | N0 | 25 (66) | 44 (29) |
|  | N1 | 8 (21) | 110 (71) |
|  | Nx | 5 (13) | 0 (0) |
| cM stage, n (%) |  |  |  |
|  | M0 | 36 (95) | 149 (96) |
|  | M1 | 2 (5) | 4 (3) |
|  | Mx | 0 (0) | 1 (1) |
| R status |  |  |  |
|  | R0 | 21 (55) | 49 (32) |
|  | R1 | 16 (42) | 67 (43) |
|  | Rx | 1 (3) | 38 (25) |

<sup>1</sup> Data available for 84/154 patients from IHC cohort
<sup>2</sup> Data available for 153/154 patients from IHC cohort

### 2.2 TMA construction

TMAs were constructed using 1 mm diameter cores punched from each formalin-fixed paraffin-embedded tissue blocks. TMAs for both DSP and IHC cohorts were retrieved from the tissue bank of the National Center for Tumor Diseases Heidelberg. The IHC cohort primarily comprised patients with high-risk PCa (11, 22). For each patient, we separated tissues into Gleason 3 and Gleason 4 categories, and, when available, also collected tissues from those with Gleason 5.

### 2.3 Sample preparation

The GeoMx® DSP platform (NanoString Technologies, Seattle, WA, USA) was used for the spatial analysis of PCa TMAs, and the TMA cores were prepared as described previously (8, 23). Briefly, slides were deparaffinized in staining jars by incubation for 3 x 5 min in xylene followed by rehydration for 2 x 5 min in 100% ethanol, 2 x 5 min in 95% ethanol and 2 x 5 min in deionized water. Antigen retrieval was performed in 1X citrate buffer (pH 6) for 15 min in a pressure cooker at high temperature and high pressure. The slides were then washed in 1X TBS-T buffer, and the tissue sections were blocked with Buffer W for 1h in a humidity chamber at room temperature (RT) before incubation with a mixture of the barcoded antibodies and morphology markers overnight at 4 °C. In this study, the NanoString barcoded antibody panels consisted of the human protein Immune cell profiling, PI3K/AKT and MAPK signaling, and cell death modules (**Supplementary Table 1**). CD45 was used to identify T-cells, panCK to label epithelial cells, and SYTO13 to stain cell nuclei, serving as morphological markers for the visualization of the tissue architecture using the NanoString Solid Tumor TME Morphology Kit. After antibody staining, slides were washed 3 × 10 min in 1X TBS-T and post-fixed in 4% paraformaldehyde at RT for 30 min, followed by 2 × 5 min washes in 1X TBS-T. Nuclei were stained with 500 nM SYTO13 for 15 min at RT and rinsed with 1X T-TBS before loading into a GeoMx® instrument (v.2.4.2.2).

### 2.4 Digital Spatial Profiling

Slides were scanned with the GeoMx instrument and regions of interest (ROIs) were selected by an experienced pathologist (S.D.). To aid in selecting ROIs, consecutive hematoxylin- and eosin-stained sections were examined simultaneously under a microscope. Each ROI was illuminated with UV light and cleaved barcodes were collected and hybridized with fluorescent probes for 16 h at 67 °C. Hybridized probes were then processed using the nCounter® MAX/FLEX Prep Station (v4.1.0.1) and counted using the nCounter® Digital Analyzer (v4.0.0.3). Quality control measures were implemented to exclude low-quality samples from the profiling analysis, and counts were normalized to housekeeping proteins GAPDH and S6. Visualization of the data was achieved using the GeoMx® DSP analysis suite and R (v.4.2.3).

### 2.5 Immunohistochemical staining

TMA cores were stained for p-JNK expression and prepared, as described previously (23). Heat-mediated antigen retrieval was performed using antigen retrieval solution (Dako, Glostrup, Denmark) and slides were blocked in goat serum. Anti-p-JNK1/2 (Thr183/Tyr185, Invitrogen/ThermoFisher Scientific, Waltham, MA, USA) was used as a primary antibody. Biotinylated anti-rabbit secondary antibody (ab97049, 1:200; Abcam) and streptavidin-peroxidase conjugate (1:1250, Merck/Sigma-Aldrich, Taufkirchen, Germany) were used to detect the primary antibody. For tissue staining and counterstaining, 3,3’-Diaminobenzidine (Abcam, Cambridge, UK) and Hematoxylin Gill I (Sigma Aldrich, St. Louis, MO, USA) were used to detect the primary antibody. TMA cores then dehydrated, and mounted with HistoMount solution (Life Technologies, Carlsbad, CA, USA). Single TMA cores were stained for p-JNK expression, according to the described immunohistochemistry protocol. P-JNK expression was assessed by a semiquantitative immunoreactive score (IRS) considering signal intensity and proportion of positively stained cells.

### 2.6 Statistical analysis and R packages

Statistical analysis of spatial protein expression data was performed using the GeoMx® DSP software (v2.4.2.2) and R (v.4.2.3). To identify statistically significant differences in protein expression between Gleason 4/5 and Gleason 3, we conducted Mann–Whitney U tests on log_2_-transformed protein expression data across the DSP cohort. *P* values were adjusted using the Benjamini-Hochberg method, with a significance threshold set at *p* < 0.1. Survival data were evaluated according to Kaplan–Meier and survival between groups was compared using the log-rank test (*p* < 0.1). R packages used for data visualization were ggplot2 (3.5.1), pheatmap (1.0.12) and survminer (0.4.9).

## 3 Results

### 3.1 DSP of protein expression in PCa patients

To characterize the spatial distribution of tumor cells in PCa, we constructed TMAs using samples from various anatomic sites within the prostate gland of 38 PCa patients (**Figure 1**). These samples were selected to represent tumor heterogeneity, incorporating regions with different Gleason scores. We selected and analyzed a total of 94 ROIs (**Supplementary Table S2**) to profile the expression of 44 proteins, including key components of the PI3K/AKT, MAPK, and cell death signaling pathways as well as immune cell markers. Due to differences in staining quality and tissue characteristics, we were able to analyze ROIs containing Gleason 3 and 4 patterns from 21 patients. Two patients had ROIs with Gleason 3 and 5, but missing Gleason 4 pattern. Additionally, 11 patients had ROIs containing either Gleason 3 or 4 pattern and four patients had ROIs corresponding to all three Gleason patterns (3, 4, and 5) (**Supplementary Table S2**). An unsupervised hierarchical clustering was performed to further explore the expression patterns. This analysis revealed significant variability in protein expression and pathway activity between Gleason 3 pattern (indolent disease) and Gleason 4 and 5 pattern (aggressive PCa) as shown in **Figure 2**.

**Figure 1.**
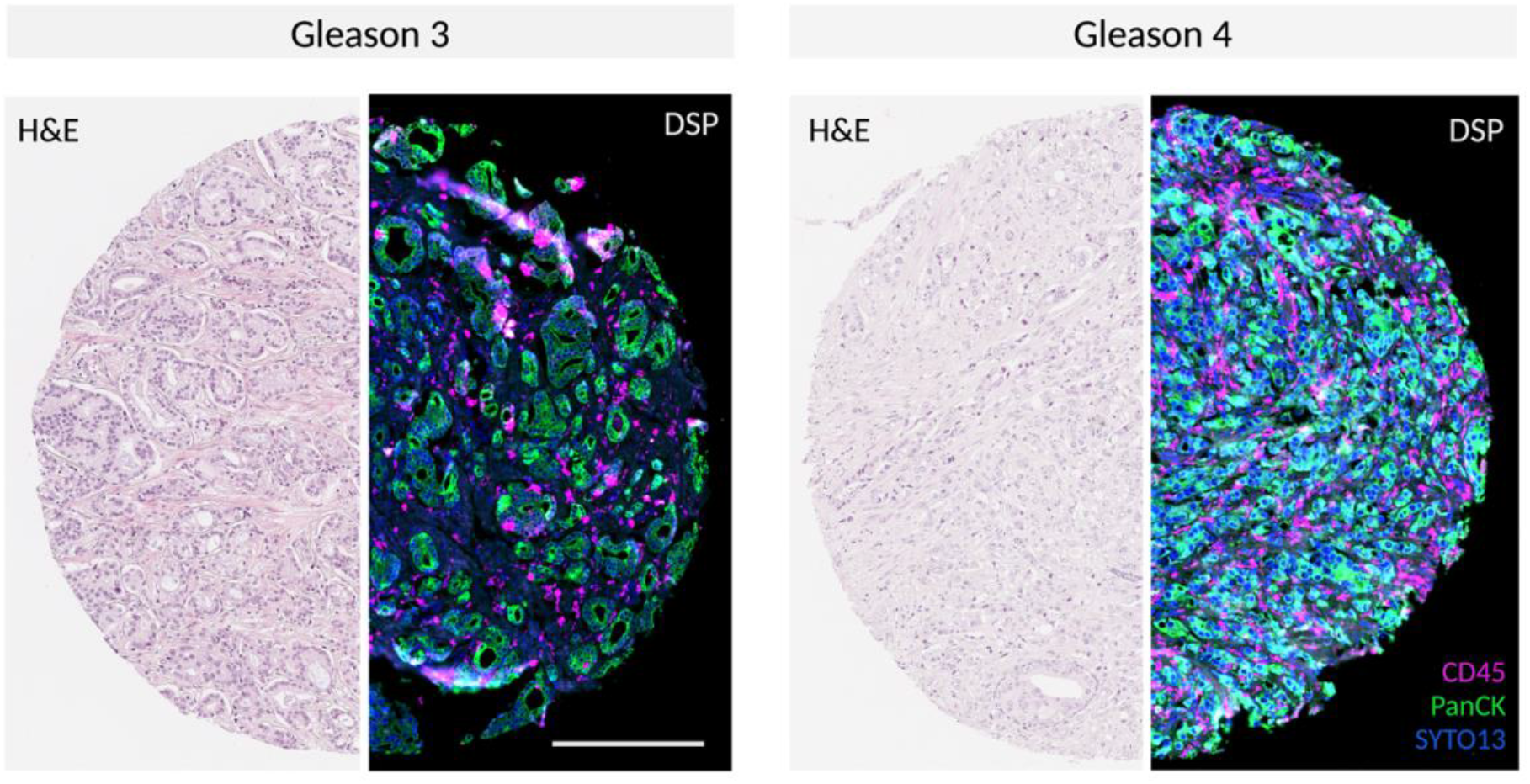
Representative GeoMx® DSP scan of PCa specimen Gleason 3 and 4 TMA cores following staining with morphology markers SYTO13, CD45 and PanCK. Scale bar = 250 µm

**Figure 2.**
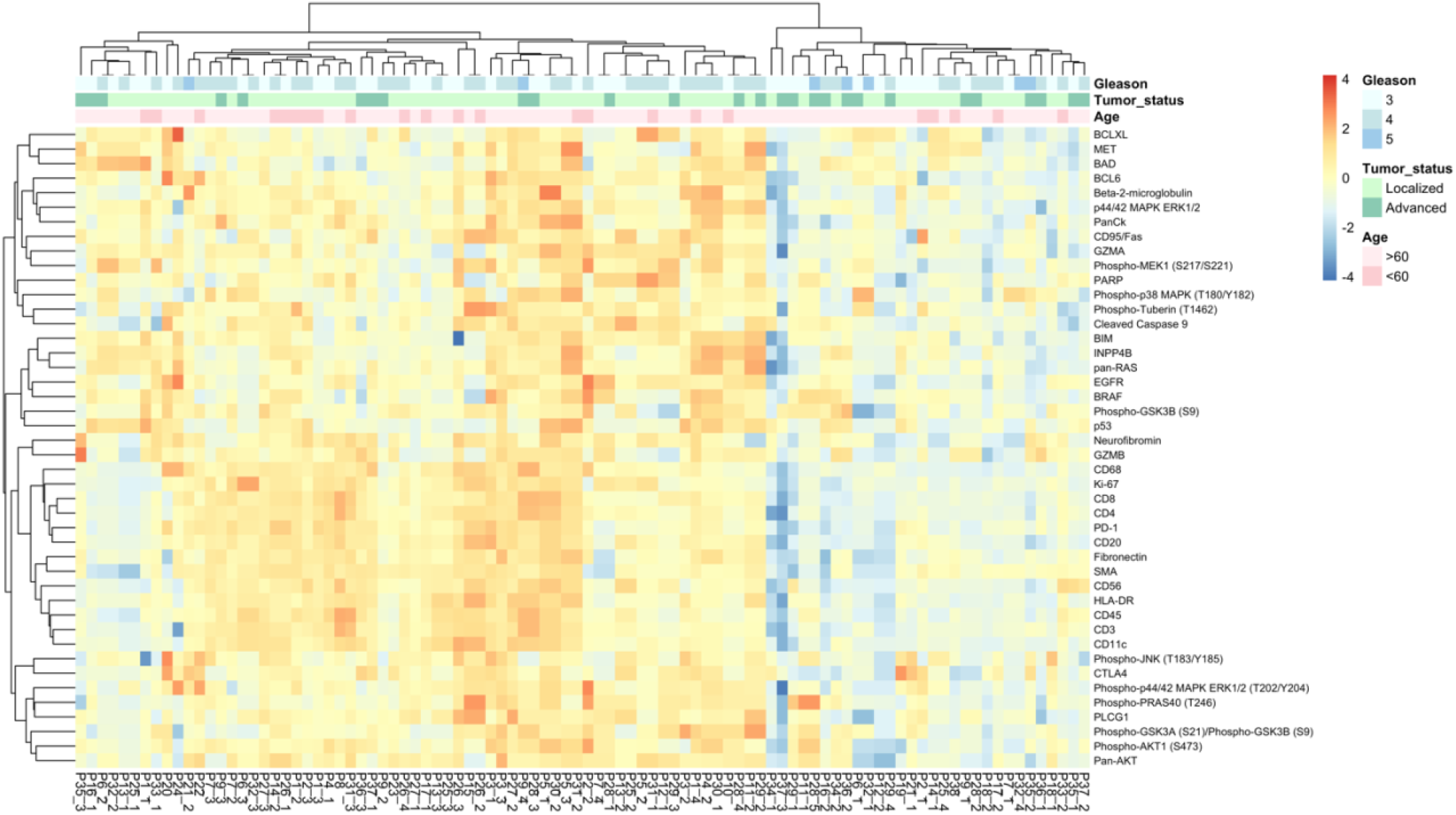
Unsupervised clustering and heatmap of DSP protein expression data obtained from 94 Gleason grade ROIs from 38 PCa patients. Data normalized to house keeper GAPDH & S6.

### 3.2 DSP identified Phospho-JNK and Ki-67 as biomarkers for prostate tumor risk stratification

The analysis of 94 ROIs derived from immune cell profiling, PI3K/AKT, MAPK signaling pathways, and cell death modules revealed the upregulation of several proteins when comparing Gleason 4 and 5 to Gleason 3 samples. While all target proteins were found to be upregulated, p-JNK and Ki-67 were the only significantly upregulated targets (adjusted *p* < 0.1) in Gleason 4 and 5 PCa compared to Gleason 3 PCa (**Figure 3A**).

**Figure 3.**
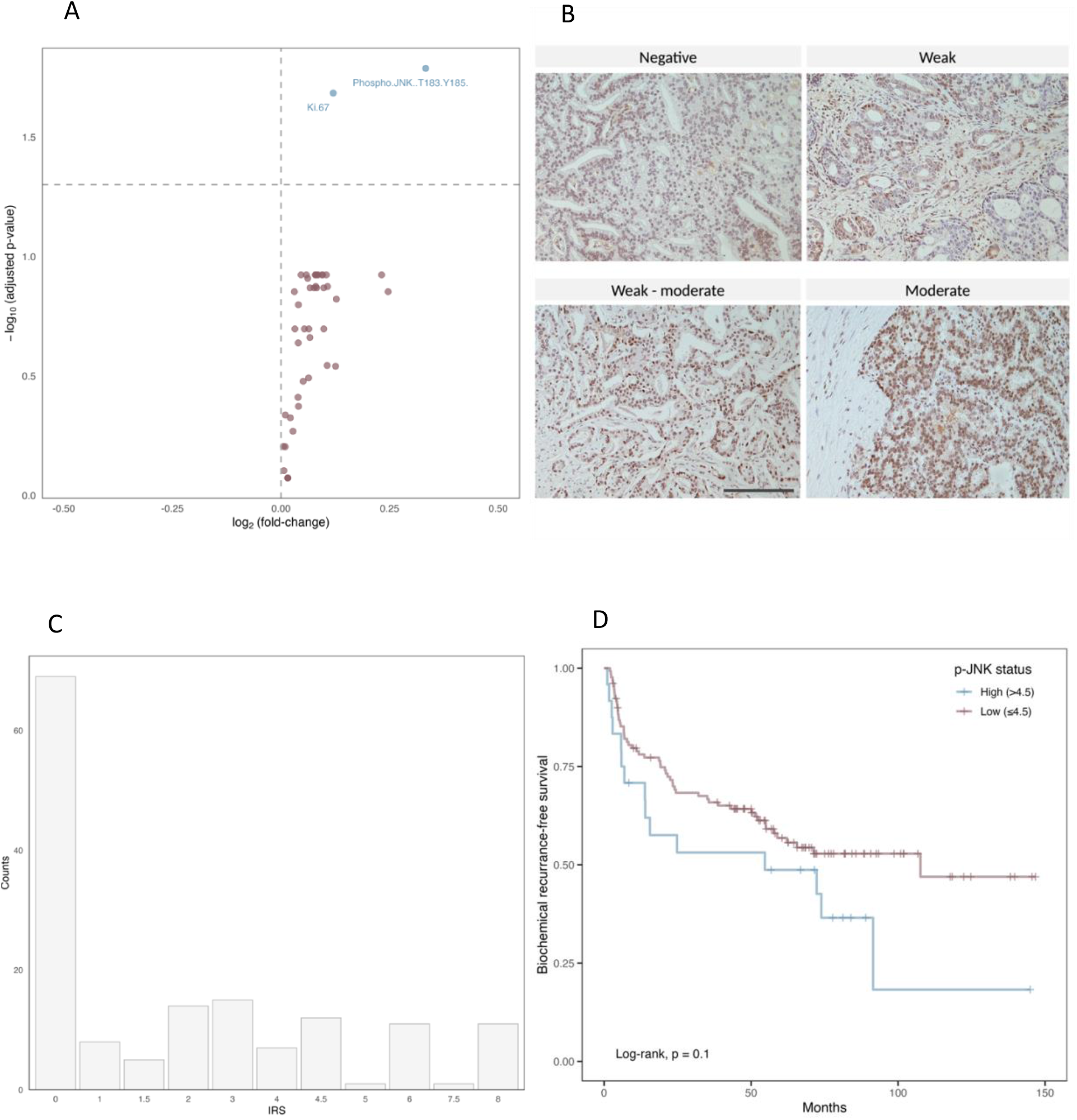
**A.** Volcano plot of 44 differentially expressed target proteins in 94 ROIs comparing Gleason 3 to Gleason 4 and 5. The vertical dotted lines represent the log_2_(fold change) of 0.5 and − 0.5. The horizontal dotted line shows the adjusted *p*-value (Benjamini-Hochberg) < 0.1. Significantly upregulated proteins are highlighted in blue. **Figure 3B**. A representative patient in the IHC cohort stained with p-JNK (Thr183/Tyr185). The staining intensity was scored on a scale of negative, weak, weak-moderate and moderate and used to calculate the IRS of p-JNK. Scale bar = 100 µm **Figure 3C**. Histogram showing the distribution of p-JNK IRS in the validation cohort of 154 patients. **Figure 3D**. Kaplan–Meier curve for 154 patients of the validation cohort, stratified into patients with high (> 4.5; n = 24) or low (≤ 4.5; n = 130) p-JNK IRS.

### 3.3 Phospho-JNK expression correlated with a shorter time to biochemical recurrence

To further investigate the role of JNK phosphorylation in tumor progression, we conducted a validation cohort analysis using TMAs of 154 PCa patients with long-term follow-up data (**Table 1**). The validation cohort was stained for p-JNK expression using IHC. For each TMA core, the p-JNK immunoreactive score (IRS) was calculated based on two components: the intensity of p-JNK staining and the fraction of positively stained cells. The staining intensity was scored on a scale of 0 to 2.5, where 0 represents negative, 1 indicates weak staining, 1.5 signifies weak to moderate staining, 2 denotes moderate staining and 2.5 corresponds to moderate to strong staining (**Figure 3B**). The proportion of positive cells was defined as follows: negative staining was scored as 0, below 10% of stained cells was scored as 1, 10–50% was scored as 2, 50% to 90% was scored as 3, and above 90% was scored as 4. The IRS was then calculated by multiplying the staining intensity by the proportion of positive cells. The highest IRS of all evaluable TMA cores per patient (TMA cores per patient [median, range] = 3, 1-6) was considered the IRS of an individual patient and the p-JNK IRS distribution ranged from 0 to 8 with a mean of 3.9 (**Figure 3C**). When patients were stratified into two groups according to p-JNK IRS, Kaplan-Meier analysis showed that patients with high p-JNK expression (p-JNK IRS > 4.5) tended to have a shorter time to biochemical recurrence (*p* = 0.1; **Figure 3D**). Therefore, these results suggest that phosphorylation of JNK is associated with aggressive PCa and a more unfavorable progression free survival in PCa.

## 4 Discussion

Heterogeneous diseases such as PCa pose significant challenges for risk stratification and clinical decision-making in patients. Addressing this challenge requires advanced diagnostic techniques to assess multiple tumor regions accurately, classify PCa subtypes, and identify biomarkers associated with aggressive disease (24). Emerging technologies such as DSP offer promising opportunities to better characterize intratumoral heterogeneity, particularly in FFPE slides, which are the standard method for storing tumor tissue (8, 25). DSP is particularly well-suited for comparing different regions within a tumor, as it enables precise selection of specific areas of interest, offering a notable advantage in spatially heterogenous tumors (11, 23, 26).

In this study, we utilized two independent cohorts of PCa patients: one cohort (n=38) to identify biomarkers for PCa aggressiveness via DSP and a larger validation cohort (n=154) with available follow-up data to correlate these markers to biochemical recurrence using IHC. Through the analysis of the spatial composition in TMA of 38 PCa patients, we demonstrated that PCa with varying Gleason scores exhibited distinct expression patterns for two key proteins, Ki-67 and p-JNK, when comparing Gleason 3 to Gleason 4 and 5 PCa. Considering that Gleason 3 tumors represent a less aggressive form of PCa, whereas Gleason 4 and 5 tumors are associated with more aggressive disease, the differential expression of Ki-67 and p-JNK suggests that these proteins may play a role in promoting tumor aggressiveness. Ki-67, a marker of cellular proliferation, has been repeatedly identified as a promising prognostic biomarker for PCa, with a higher Ki-67 index correlating with worse biochemical recurrence-free survival (27). JNK, a member of the MAPK family, regulates a wide range of cellular processes (28). In this study, we further focused on p-JNK because, unlike Ki-67, its role as a biomarker for PCa aggressiveness remains less established. In our validation cohort of 154 PCa patients, immunostaining with p-JNK antibody (Thr183/Tyr185) revealed that higher p-JNK expression was associated with a shorter time to biochemical recurrence, highlighting the role of p-JNK upregulation in tumor progression.

JNK has a dual role in cancer, acting either as a tumor suppressor by inducing cell death or as a promoter of cell proliferation, depending on factors such as the type of stimuli, tissue specificity, and signal intensity (29, 30). In response to stress, JNK can initiate cell death by activating pro-apoptotic transcription factors (31). Conversely, the loss of JNK signaling can contribute to tumor formation through phosphorylating of specific signaling proteins that simulate growth-related gene expression (28). In its dual paradoxical role, JNK also promotes cell survival by downregulating FoxO1-dependent autophagy (32-34). However, prolonged JNK activation often leads to apoptosis mediated by TNFα (35). In other tumor entities, JNK pathway deficiency was shown to support HER2+-driven breast cancer (36), while elevated JNK expression was associated with worse prognosis in colorectal cancer patients (37). Increasing evidence indicates that JNK signaling is closely related to cellular senescence, a state of permanent growth arrest (38). The secretory profile of senescent cells can modify the tissue microenvironment by evading immune surveillance and creating conditions that robustly drive PCa progression (39, 40).

In PCa, JNK contributes to both apoptosis and tumor progression, reflecting its complexity in cancer pathways. Activation of JNK has been shown to increase sensitivity to chemotherapy and promote apoptosis in PCa cells, indicating its potential in sensitizing tumors to treatment (41). Similarly, inhibition of autophagy through the JNK pathway significantly enhances apoptosis in PC3 cells (42). However, JNK also promotes prostate tumor growth through interactions with the tumor microenvironment (40, 42). Higher JNK expression corelates with higher Gleason score and is associated with shorter overall and progression-free survival in patients with castration-resistant PCa (43). These findings suggest that targeting JNK pathway could be a promising therapeutic strategy for PCa, potentially improving the effectiveness of current treatments. In addition, JNK as a biomarker in early PCa could help differentiate aggressive from indolent disease, thereby improving decision-making regarding active surveillance versus radical treatment in PCa.

The limitations of this study include the small DSP cohort and a limited size of the protein panel. The focus on analyzing high-level protein expression excludes low-expressed proteins, as the panel is predominantly composed of oncogenes that are upregulated in PCa. Furthermore, obtaining this information from tissue requires a tissue biopsy, which carries potential risks and side effects for the patient.

In summary, differentially expressed proteins in aggressive versus indolent PCa can shed light on the molecular mechanisms driving the transition from a lower-grade to a higher-grade PCa. Targeting specific proteins that are upregulated in high-grade tumors could serve as potential biomarkers and open new therapeutic avenues. This approach can ultimately contribute to the field of personalized medicine by helping clinicians to tailor treatment strategies based on the molecular characteristics of the tumor.

## Data Availability

All data produced in the present work are contained in the manuscript

## Ethics approval and consent to participate

This study was conducted in accordance with the Declaration of Helsinki and approved by the Ethics Committee of the Medical Faculty of the University of Heidelberg (votes: S-130/2021 and S-864/2019).

## Availability of Data and Materials

The data presented in this study are available in the article.

## Authorship

Conceptualization, S.E., A.K., S.D., A.D. and M.G.; Methodology, S.E. and A.K.; Software, S.E., A.K., F.J. and S.B.; Validation, S.E., A.K., F.J., S.D., A.D. and M.G.; Formal Analysis, S.E. and A.K.; Investigation, S.E., A.K., A.L.R., O.L., G.K., C.S., A.S., H.S., S.D., A.D. and M.G.; Resources S.D., A.D. and M.G.; Data Curation, S.E., A.K., S.D., A.D. and M.G. Original Draft Preparation, S.E.; Review and Editing, all authors; Visualization, S.E.; Supervision, H.S., S.D., A.D. and M.G. All authors have read and agreed to the published version of the manuscript.

## Acknowledgements

192 prostate cancer samples were provided by the Tissue Bank of the NCT Heidelberg, Germany, in accordance with the regulations of the biobank and the approval of the ethics committee of Heidelberg University. In addition, we would like to thank Daniela Janscho for her expert technical support.

## Legends

**Supplementary Table 1.** Overview of Nanostring GeoMx® DSP protein panels.

**Supplementary Table 2.** Overview of histopathological information for each patient and each ROI.

