## Supplementary Table S1 for "Digital Spatial Profiling identifies phospho-JNK as a biomarker for early risk stratification of aggressive prostate cancer"

### Supplementary Table 1

| Immune Cell Profiling | MAPK Signaling | Cell Death | PI3K/AKT signaling |
|---|---|---|---|
| PD-1 | BRAF | BAD | Phospho-AKT1 (S473) |
| Pan-cytokeratin | EGFR | BCL6 | Phospho-GSK3B (S9) |
| CD68 | Phospho-JNK (T183/Y185) | BCLXL | Phospho-GSK3A (S21)/Phospho-GSK3B (S9) |
| HLA-DR | Phospho-MEK1 (S217/S221) | CD95/Fas | INPP4B |
| SMA | Phospho-p38 MAPK (T180/Y182) | GZMA | PLCG1 |
| Ki-67 | Phospho-p44/42 MAPK ERK1/2 (T202/Y204) | Cleaved Caspase 9 | Phospho-PRAS40 (T246) |
| Beta-2-microglobulin | pan-RAS | p53 | Phospho-Tuberin (T1462) |
| CD11c | Phospho-p44/42 MAPK ERK1/2 (T202/Y204) | PARP | Pan-AKT |
| CD20 | Phospho-p90 RSK (T359/S363) | BIM | MET |
| CD3 | | Neurofibromin | |
| CD4 | | | |
| CD45 | | | |
| CD56 | | | |
| CD8 | | | |
| CTLA4 | | | |
| GZMB | | | |
| PD-L1 | | | |
| Fibronectin | | | |
| GAPDH | | | |
| Histon H3 | | | |
| S6 | | | |
| Rb IgG | | | |
| Ms IgG1 | | | |
| Ms IgG2a | | | |
