## Supplementary Table S2 for "Digital Spatial Profiling identifies phospho-JNK as a biomarker for early risk stratification of aggressive prostate cancer"

| TMA | Patient_ID | Tumor_status | ROI | Gleason |
|---|---|---|---|---|
| 1 | P1 | Localized | 1 | 3 |
| | | | 2 | 4 |
| | | | 3 | 3 |
| | | | 4 | 4 |
| | P2 | Localized | 5 | 3 |
| | | | 6 | 4 |
| | | | 7 | 4 |
| | P3 | Localized | 8 | 3 |
| | | | 9 | 4 |
| | | | 10 | 4 |
| | P4 | Localized | 11 | 3 |
| | | | 12 | 4 |
| | P5 | Localized | 13 | 3 |
| | | | 14 | 4 |
| | | | 15 | 4 |
| | P6 | Advanced | 16 | 3 |
| | | | 17 | 4 |
| | | | 18 | 3 |
| | P7 | Localized | 19 | 3 |
| | | | 20 | 4 |
| | | | 21 | 4 |
| | | | 22 | 3 |
| | P8 | Localized | 23 | 4 |
| 2 | P9 | Advanced | 24 | 3 |
| | | | 25 | 4 |
| | | | 26 | 4 |
| | | | 27 | 5 |
| | P10 | Localized | 28 | 3 |
| | P11 | Localized | 29 | 3 |
| | | | 30 | 4 |
| | P12 | Localized | 31 | 4 |
| | | | 32 | 3 |
| | P13 | Localized | 33 | 4 |
| | | | 34 | 3 |
| | | | 35 | 4 |
| | P14 | Localized | 36 | 3 |
| | | | 37 | 4 |
| | P15 | Localized | 38 | 4 |
| | P16 | Advanced | 39 | 3 |
| | | | 40 | 4 |
| | P17 | Localized | 41 | 3 |
| | | | 42 | 4 |
| | P18 | Localized | 43 | 3 |
| | | | 44 | 4 |

| | | | | |
|---|---|---|---|---|
| 3 | P19 | Localized | 45 | 3 |
| | P20 | Localized | 46 | 3 |
| | P21 | Localized | 47 | 3 |
| | | | 48 | 5 |
| | P22 | Localized | 49 | 4 |
| | P23 | Localized | 50 | 4 |
| | P24 | Localized | 51 | 4 |
| 4 | P25 | Localized | 52 | 3 |
| | | | 53 | 4 |
| | | | 54 | 3 |
| | | | 55 | 4 |
| | P26 | Localized | 56 | 3 |
| | | | 57 | 4 |
| | | | 58 | 3 |
| | | | 59 | 4 |
| | P27 | Localized | 60 | 3 |
| | | | 61 | 4 |
| | | | 62 | 3 |
| | P28 | Advanced | 63 | 3 |
| | | | 64 | 4 |
| | | | 65 | 3 |
| | | | 66 | 4 |
| | | | 67 | 5 |
| | P29 | Advanced | 68 | 3 |
| | | | 69 | 4 |
| | | | 70 | 3 |
| | | | 71 | 4 |
| | P30 | Localized | 72 | 4 |
| | | | 73 | 4 |
| | P31 | Localized | 74 | 4 |
| | | | 75 | 3 |
| | | | 76 | 4 |
| | P32 | Localized | 77 | 5 |
| | | | 78 | 3 |
| | | | 79 | 4 |
| | | | 80 | 5 |
| 5 | P33 | Localized | 81 | 4 |
| | | | 82 | 4 |
| | P34 | Localized | 83 | 3 |
| | | | 84 | 4 |
| | P35 | Advanced | 85 | 3 |
| | | | 86 | 5 |
| | | | 87 | 3 |
| | P36 | Advanced | 88 | 4 |
| | | | 89 | 5 |
| | | | 90 | 3 |

| | | | | |
|---|---|---|---|---|
| | P37 | Advanced | 91 | 3 |
| | | | 92 | 4 |
| | | | 93 | 3 |
| | P38 | Localized | 94 | 4 |
